# Physical and psychiatric comorbidity among patients with severe mental illness as seen in Uganda

**DOI:** 10.1101/2020.11.04.20225813

**Authors:** Richard Stephen Mpango, Wilber Ssembajjwe, Godfrey Zari Rukundo, Carol Birungi, Allan Kalungi, Kenneth D. Gadow, Vikram Patel, Moffat Nyirenda, Eugene Kinyanda

**Affiliations:** Mental Health Section, MRC/UVRI and LSHTM Uganda Research Unit & Senior Wellcome Trust Fellowship, Entebbe, Uganda; Butabika National Psychiatric Hospital, Kampala, Uganda; Statistical Section, MRC/UVRI and LSHTM Uganda Research Unit, Entebbe, Uganda; Department of Psychiatry, Mbarara University of Science and Technology, Uganda; Department of Psychiatry, Stony Brook University, Stony Brook, New York; Department of Global Health and Social Medicine, Harvard Medical School, Massachusetts, USA; Department of Psychiatry, College of Health Sciences, Makerere University, Kampala, Uganda; Global Non-Communicable Diseases (NCD) Section, MRC/UVRI and LSHTM Uganda Research Unit, Entebbe, Uganda; Deartment of Mental Health, School of Health Sciences, Soroti University, Soroti, Uganda

## Abstract

This study established the prevalence of physical and psychiatric comorbidity and associated risk factors among 1,201 out-patients with severe mental illness (SMI) attending Butabika and Masaka hospitals in Uganda. Participants completed an assessment battery; structured, standardized and locally translated instruments. SMIs were established using the MINI International Neuropsychiatric Interview version 7.2. We used logistic regression to determine the association between physical and psychiatric comorbidity and risk factors. Prevalence of physical and psychiatric comorbidity was 13.1 %. Childhood sexual abuse (aOR 1.06, 95% CI 1.03 -1.10, P=0.001), sexual abuse in adulthood (aOR 2.22, 95% CI 1.60 - 3.08, P<0.001), childhood physical abuse (aOR 1.07, 95% CI 1.03 - 1.10, P<0.001) and physical abuse in adulthood (aOR 1.69, 95% CI 1.30 - 2.20, P<0.001) were associated with an increased risk of having comorbid psychiatric and physical disorders. Emerging healthcare models in Uganda should optimise care for people with physical and psychiatric comorbidity.

## Introduction

Mental ill health is a major cause of morbidity and mortality globally, with low- and middle-income countries (LMIC) experiencing a rapidly increasing prevalence of morbidity (1). Patients with severe mental illness (SMI) such as schizophrenia, depression and bipolar affective disorder die younger compared to the general population (2-4). However, most of the increased mortality among people with SMI is attributed to comorbidity with other non-communicable diseases (NCDs) (2, 3, 5). Patients with SMI experience a range of chronic physical health problems which may interfere with quality of life, increase health seeking behaviour and contribute to poor treatment outcomes (6). In turn, mental illness contributes to the NCD risks and outcomes, through adoption of harmful behaviours (such as sedentariness, smoking or excessive alcohol intake) which may affect treatment adherence and retention in care (1). Secondly, some medications for SMI, particularly the second-generation antipsychotics are associated with increased risk of obesity, diabetes and metabolic syndrome (7). Lastly, there is a tendency for comorbidity among the severe mental illnesses; there is an increased risk for the development of depression among patients with schizophrenia (8-10); the overlap in the symptoms and genetic risk factors between psychiatric disorders suggests a common etiological mechanism (8).

Physical and psychiatric comorbidity has become increasingly important among people with SMI receiving mental health services at hospitals due to several risk factors (11). Patients with comorbidity are therefore generally thought to have higher rates of healthcare utilisation and poorer outcomes, in part because they are at risk of receiving suboptimal care for the co-existing conditions (12, 13). On the other hand, comorbidity may provide an opportunity for patients to receive care beyond the index condition (14, 15). This is best illustrated in HIV care in sub-Saharan Africa where the control of hypertension and other NCDs is better among HIV positive than HIV negative individuals, perhaps because individuals living with HIV have more regular and intensive contact with the healthcare system (16, 17).

Although comorbidity is of worldwide concern, the majority of evidence to date comes from high-income countries, with only a few studies from LMIC, and fewer from sub-Saharan Africa (1, 18). These limited data suggest a high and increasing burden of comorbidity (18, 19), affecting relatively young people (20). Detection and management of mental and physical comorbidity will pose a particular challenge for LMIC healthcare systems, which historically have separated services for mental and physical health (1, 21). The purpose of our study was to establish the prevalence of comorbidity, associated risk factors and negative outcomes among patients with severe mental illness (SMI) in rural and urban Uganda.

## Results

### Characteristics of study participants

Table 1, of the 1201 participants enrolled into this study, 39.7% were between 35 and 49 years and 32.1% were between 25 and 34 years; mean age (with standard deviation) was 37.6(11.7). The urban and rural study sites contributed 58% and 42% of participants respectively. About a third each of the participants were single (38.7%) or married (32.0%) and a quarter (24.6%) separated/divorced, with 4.7% widowed. Males and females enrolled in the study were 46% and 54% respectively. Christians were 81% and Muslims were 18%. More than half (57.2%) of the respondents had at least seven years of formal education. A third (32.9%) had suffered childhood physical abuse and a quarter (24.7%) childhood sexual abuse. Physical abuse in adulthood was reported by 34.1%, while sexual abuse in adulthood was reported by 21.9%. About two thirds (61.0%) reported a family history of psychiatric disorder.

### Pattern of psychiatric and physical comorbidity

Table 2, bipolar affective disorder was the most prevalent (66.4%) primary psychiatric diagnoses followed by schizophrenia (26.6%) and recurrent major depressive disorder (7.0%). Psychosis was the most prevalent (22.2%) current psychiatric episode followed by depressive episode (13.5%) and manic episode (6.2%). Psychiatric comorbidity was reported by 9.1% of respondents, while physical disorder/problems comorbidity was reported by 42.6%. The specific comorbid physical disorder/problems were, 27.1% hypertension, 13.8% obesity, 8.2% HIV/AIDS and 4.8% syphilis.

**Table 2:** Pattern of psychiatric and physical comorbidities among respondents

| **Factor** | **Level** | **N=1201** | **Prevalence (95% CI)** |
| --- | --- | --- | --- |
| **Primary psychiatric diagnosis** |  |  |  |
| Schizophrenia | Yes | 320 | 26.6% |
| Bipolar affective disorder | Yes | 797 | 66.4% |
| Recurrent Major depressive disorder | Yes | 84 | 7.0% |
| **Comorbid physical disorder/ problem** |  |  |  |
| HIV/AIDS | Positive | 87/1057 | 8.2% (6.7% - 10.0%) |
| Syphilis | Positive | 53/1095 | 4.8% (3.7% - 6.3%) |
| Hypertension | Sys≥140 dia≥90 | 326/1201 | 27.1% (24.7% - 29.7%) |
| Obesity | BMI≥30 | 165/1193 | 13.8% (11.9% - 15.9%) |
| Had at least one comorbid physical disorder/problem | Yes | 512/1201 | 42.6 % ( 39.8% - 45.4%) |
| Had at least two comorbid physical disorder/problem | Yes | 109/1201 | 9.1 % ( 7.5% - 10.8%) |
| **Current psychiatric episode & comorbidity** |  |  |  |
| Manic episode | Yes | 74/1196 | 6.2 % ( 4.8% - 7.6%) |
| Psychotic episode | Yes | 263/1187 | 22.2 % ( 20.0% - 24.8%) |
| Depressive episode | Yes | 160/1189 | 13.5 % ( 11.5% - 15.4%) |
| Did not meet criteria for at least one current psychiatric disorder episode | Yes | 831/1201 | 69.1 % ( 66.5% - 71.7%) |
| At least one physical and one psychiatric comorbidity | Yes | 157/1201 | 13.1 % ( 11.3% - 15.1%) |
| Had at least two current psychiatric disorder episodes | Yes | 109/1201 | 9.1 % ( 7.5% - 10.8%) |

### Factors associated with psychiatric and physical comorbidity

Table 3 and 4, risk factors for psychiatric comorbidity were: age category of 25-34 years compared to those aged between 18-24 years, increasing mental health stigma, childhood physical abuse, childhood sexual abuse, physical abuse in adulthood, sexual abuse in adulthood and use of mood stabilisers. Protective factors against psychiatric comorbidity included rural residence (compared to urban residence) and increasing social support. Risk factors of HIV comorbidity were female gender and being widowed and separated (compared to being married). Being Moslem (compared to being Christian), increasing socio-economic status and having a past psychotic episode were protective against HIV.

The risk factor for syphilis comorbidity were increasing age, being widowed and separated and alcohol use. The risk factors of hypertension comorbidity were increasing age and alcohol use, while rural residence was protective against hypertension. Risk factors for obesity were female gender, increasing age, increasing socio-economic status, use of mood stabilisers, problem alcohol drinking, use of marijuana and use of Khat, while staying in a rural residence was protective.

### Association between psychiatric and physical comorbidity with negative behavioural and clinical outcomes

Table 5, only psychiatric comorbidity was associated with the negative outcomes of life time attempted suicide and risky sexual behaviour. None of the physical disorders/problems was associated with the investigated negative behavioural and clinical outcomes.

## Discussion

This study demonstrates a high prevalence of physical and psychiatric comorbidity, associated risk factors and negative outcomes among patients with severe mental illness (SMI) attending out-patients’ departments (OPD) at Butabika and Masaka hospitals in Uganda. According to this study, comorbidity between psychiatric disorders and a physical disorders was reported by 13.1 % of the respondents, while a meta-analysis established the pooled prevalence of 36.6% for psychiatric disorders in patients with chronic physical diseases (22). Obviously, difference between prevalence rates established by this study (cross-sectional at baseline) and the meta-analysis by Daré et al (22) relate to the different study designs. Similarly, the prevalence of psychiatric disorders in people with chronic physical diseases living in developing and emerging countries is comparable to those in developed countries (22). Importantly, the relationship between medical and psychiatric illness involves multiple factors (23), and happens to be bi-directional; physical illness can cause the mental illness but also the mental illness can cause the physical illness (24). The co-occurrence of psychiatric and physical disorders is supported by the mind-body interaction (substance dualism) suggested by Descartes (1641) as described by Nadler S & Morris K (1997) (25).

The prevalence of psychiatric comorbidity (presence of at least two current psychiatric disorder episodes) reported in this study was 9.1%, which was much lower than the 24% of respondents who met the criteria for two psychiatric disorders in a study about the prevalence of psychiatric disorders on the general wards of Mbarara regional referral hospital in Southwestern Uganda; both studies used the Mini International Neuropsychiatric Interview (MINI) to determine specific psychiatric diagnoses but this study interviewed participants attending out-patient departments (OPDs), while the study by Rukundo et al. (2013)(23) interviewed admitted patients. The degree of psychiatric morbidity happens to be directly related to indicators of family adversity, physical abuse, other psychosocial variables (26), and interpersonal adversity experienced since childhood (27). Similarly, poverty, homelessness, substance use, and smoking all augment the risk of both physical and psychiatric illness (24).

Psychiatric comorbidities are so common and might be integral to schizophrenia (28). Commonly, the symptomatology of schizophrenia over-shadows other psychiatric disorders thus it is difficult to determine primacy but the alternative approach adopted by Diagnostic and Statistical Manual of Mental Disorders (DSM-IV R) is to consider these symptoms as part of another axis I diagnosis that occurs alongside schizophrenia (29). Possibly, depressive symptoms in schizophrenia are associated with antipsychotic medications that produce neurological side effects like Parkinsonism (particularly bradykinesia, diminution of affective expression, masked facies, and verbal delays) and akathitic restlessness that may be confused with the psychomotor retardation or agitation of depression (28). Antipsychotic drugs may also produce a primary dysphoria, possibly due to dopamine blockade in reward pathways, and it has even been suggested that these drugs are innately depressogenic (28). Similarly, the classic construct of depression in schizophrenia is that of post-psychotic depression (PPD), defined in an appendix of the DSM-IV R (29), as a major depressive episode that is superimposed on, and occurs only during the residual phase of schizophrenia. Traditionally, depression in schizophrenia has been formulated as a psychological reaction to loss or to the psychological trauma of the psychotic episode (28). Arguably, depression is a reaction to psychosis, or represents an unmasking effect of the depression as the psychosis remits (28). Similarly, there is a clear substantial comorbidity index between bipolar disorder and schizophrenia (30, 31).

On the individual physical comorbidities, this study reported a rate for HIV co-morbidity of 8.2% against a national general population prevalence of HIV of 5.7%(32). Two earlier Ugandan studies reported higher rates of HIV co-morbidity among patients with SMI, a rate of 18.4% in 2011(33) and a rate of 11.3% in 2013 (34), this was against a national general population prevalence of HIV of 7.3% (35, 36). In all these three studies, the rate of HIV among patients with SMI was generally higher than that in the general population, suggesting that patients with SMI were at increased vulnerability to HIV compared to the general population. In this study, the least reported physical comorbidity, was syphilis at a rate of 4.8%, against a Uganda national prevalence rate of 2.9% in 2016 among women attending antenatal care (37). The rate of syphilis comorbidity in this Ugandan study was twice that reported in a Brazilian study (1.1%) a country where the general prevalence of syphilis is reported at 7.7% among women attending antenatal care (38). Both vulnerability to HIV and syphilis which are predominantly transmitted through heterosexual relationships in the SMI context in Uganda are underlined by a common vulnerability to risky sexual behaviour and sex violence which Lundberg and colleagues (2015) in their Ugandan study reported to be more prevalent in SMI than in the general population (39).

In this study, the most prevalent comorbidity reported was hypertension (27.1%), this was against a Uganda general population rate of hypertension of 26.4% (40). This rate of comorbid hypertension in SMI is much higher than that reported by two earlier UK studies, a rate of 19% in 2014 (41) and a rate of 18.3% in 2015 (42) against a general population rate of hypertension in the UK of 26.2% in 2017 (43).

The prevalence of comorbid obesity in this study was 13.4%, this is against a Uganda general population rate of obesity of 7 % (44). A study among patients with SMI in the USA published in 2010 reported a rate of obesity of 52% (45), against a general population rate of 35.7% in the USA in 2009–2010 (46). Factors underlying the association between SMI and the cardiovascular risk factors of hypertension and obesity include a variety of lifestyle factors such as smoking, lack of physical activity, and poor diet (45).

The presence of psychiatric disorders in medically ill patients increases the cost of health care due to repeated ineffective use of services, increased length of hospital stay, hospitalization rates and mortality (23). It is for this reason that the World Health Organization recommended the integration of mental health services into general health care services (47), in order to effectively and efficiently respond to the psychiatric and physical comorbidity (22).

### Pattern of psychiatric and physical comorbidities among patients with severe mental illness in Uganda

The clustering of psychiatric disorders (with participants having two or more current psychiatric disorders), relates to previous literature which suggests that chronic physical, infectious and mental health conditions commonly co-exist (1, 48). A previous study established that psychiatric conditions (such as schizophrenia, depression, bipolar affective disorders, etc.) cluster with a range of physical NCDs as well as with chronic infections such as HIV (12). In addition to ageing, increased prevalence of comorbidity among people in low- and middle-income countries (LMICs) relates to a growing prevalence of NCDs (such as an increase in obesity and physical inactivity) in addition to the well-known burden of infectious diseases like HIV/AIDS (48). This change in condition patterns reveals the changing lifestyle, cultural behaviours, changing environmental exposures, urbanization, and healthcare-related advances that contribute to an increased prevalence of chronic conditions (49, 50). The relationship between mental health and physical conditions appears to be bidirectional and may arise due to shared biological factors, or mediated by various lifestyle and treatment specific factors (12). Similarly, the clustering of physical comorbidity with index psychiatric disorders was high because comorbidity is generally higher among mentally sick people who happen to be more vulnerable, socioeconomically disadvantaged, often have a lower capacity to access healthcare and deal with the burden of ill health (48, 51). Relatedly, some studies have found that those with comorbidity (especially those on antipsychotic treatment) are at particular risk of adverse drug events (52) and are at risk of receiving suboptimal care for co-existing physical conditions (12, 13). Conditions appear to cluster in many LMICs in which HIV infection is common and the use of ART widespread (49). It has been established that use of antiretroviral therapies (ART) for the treatment of HIV has been associated with insulin resistance, elevated blood lipids, and central fat accumulation, each of which can ultimately contribute to the development of type 2 diabetes and cardiovascular diseases (48). Depression has also been shown to increase the risk of morbidity and mortality in populations with type 2 diabetes (53). Some psychotropic medications, most notably antipsychotics, have also been reported to be associated with increased risk of several physical conditions including obesity, diabetes, cardiovascular diseases, and haematological diseases (54, 55). It has also been suggested that chronic inflammation and oxidative stress may underlie a number of other chronic conditions and cancers, and could therefore also contribute to several comorbidity clusters (56-59). Given the common co-occurrence of mental and physical health conditions in the context of comorbidity, there is a need for better evidence about causality and confounding in order to identify the exacerbatory effects of medications on those with comorbidity.

### Association between socio-demographic factors and comorbid psychiatric and physical disorders

Relatedly, a growing body of evidence suggests that physical illnesses and multimorbidities are significantly more prevalent in the population of psychiatric patients than in the general population because mental disorders are associated with an increased risk of a wide range of chronic physical illnesses and comorbidity (60). According to this study, staying at Masaka was a protective by 23% against psychiatric comorbidity and the findings are in agreement with results from a previous study which suggests that living in rural areas is significantly associated with lower risk of reporting severe mental illnesses and happens to be associated with better overall mental health (61). Result from this study indicate that the age category of >=50 was protective by 39% against psychiatric comorbidity, yet other studies suggest that increasing age is associated with increased comorbidity (62, 63); possible explanation could be that increased social support (often attained with increasing age) buffers psychological distress and happens to be protective against psychiatric comorbidity (64).

Based upon results from this study, being a female increased the risk of having HIV by almost three-times and the findings happen to be in agreement with a previous study which suggests that gender disparities put women at an increased risk of HIV (65, 66); gender inequity is manifested in the social and economic burden women carry in relation to men (66). Related to findings from this study, a recent Ugandan report (2017) indicated that in the age category of 15-64years, HIV was more prevalent among females (7.6%) as compared to males (4.7%)(67). Similarly, higher HIV prevalence rates among females as compared to males could be due to other socio-cultural factors (66, 68). Being a moslem was protective by 69% against HIV (aOR 0.31, 95% CI 0.13 - 0.72, *P=0*.*007*); the findings from this study are in agreement with previous studies which suggest the efficacy of male circumcision (mostly practiced by moslems) in reducing the risk of acquiring HIV and some sexually transmitted infections (69-71). Increase in social economic status (indicated by ses score) was a protective by 13% against HIV; these finding are in agreement with a previous study which suggested an association between social economic status and HIV (68, 72). Marital status (being single) was protective by 28% against HIV, thus happens to be in agreement with findings from a study undertaken in Kenya which established that the risk of acquiring HIV was significantly associated with being married, divorced/separated/widowed (73).

The age category of >=50 had a five-fold increased risk of having syphilis and the results happen to be in agreement with a study undertaken in Brazil (developing country) which suggests that the prevalence of syphilis among people with SMI ranges between 1.1% to 7.6% (74). Similar to findings from this study, a recent survey in Uganda revealed that the prevalence of active syphilis infection among people with HIV in the age category of 15-64 years was similar (2%) among men and women (67). Similarly, marital status (being single) was protective by 19% against syphilis ; a related study established that concurrent sexual partners are associated with an increased prevalence of syphilis (75).

Results from this study indicates that staying at Masaka was a protective by 36% against hypertension, which happens to be in agreement with results from a recent survey with findings indicating that hypertension is mostly common among people living in the urban compared to those in rural areas due to the differences in lifestyles (76, 77). The age category of >=50 had four-fold increased risk of hypertension and these findings happen to agree with results from a study which suggests that the increasing age is a risk factor for hypertension among people with SMI (78).

Staying at Masaka was a protective by 57% against obesity which happens to be in agreement with results from a recent survey with findings indicating that obesity is mostly common among people living in the urban compared to those in rural areas due to the differences in lifestyles (76, 77). Similarly, being female had a five-fold increased risk of having obesity; a previous study suggested that sex differences in gene expression are related to antipsychotic induced weight gain (AIWG); biological processes are involved in AIWG and provide additional evidence of the genetic links between weight gain and antipsychotic treatment (79). Findings from this study are in agreement with a previous study which suggests that hypertension and obesity happens to be more common among women than men (77) due to biological, socio economic factors and gender differences in the roles. Related to findings from this study, recent surveys indicated that females had a higher prevalence of obesity than males (females 7.5% ; males 1.8%) (40, 76). According to this study, people with SMI in the age category of >=50 had a four-fold increased risk of obesity; emerging evidence suggests that the most significant cause of weight gain relates to the metabolic side effects of antipsychotic medication (80). Probably, the increased risk of obesity among people with SMI in the age category of >=50 could be a result of ageing, years spent on antipsychotic treatment, coupled with the sedentary behaviour (1). Interestingly, increase in social economic status was a risk factor for obesity and these findings happen to be in agreement with previous studies undertaken in the region (81, 82).

### Psychosocial and psychiatric factors associated with comorbid psychiatric and physical disorders

According to this study, social support was protective by 2% against psychiatric comorbidity since social support acts as a buffer against psychological distress (64), thus a supportive social environment increases the survival of people with chronic illnesses (83). Mental health stigma was a risk factor for psychiatric comorbidity which rhymes with findings from other studies which suggest that social stigma is common among people with psychiatric comorbidity and increases as more insight is regained (84) ; social stigma is largely related to the double burden of symptoms and pressure to undertake treatment for both their mental and physical diseases, questionable prognosis and the likely associated negative outcomes (85). History of childhood physical abuse was a risk factor for psychiatric comorbidity; findings from this study are in agreement with previous studies which suggests that there is an association between self-reported childhood difficulties and later comorbidity in adulthood (86, 87). The high level of childhood physical abuse relates to the disciplinary measures and corporal punishment widely used in Uganda as a form of parenting, with much uncertainty as to whether they are beneficial for discipline or risk factors for mental Illness (88, 89); childhood physical abuse is significantly associated with later mental distress (90). History of childhood sexual abuse was a risk factor for psychiatric comorbidity; findings from this study happen to be in agreement with previous studies (86, 87). Previous studies in Uganda have reported high levels of sexual abuse commonly experienced by girls (89, 91), which happens to be a risk factor for poor mental health (91). Previous studies established that childhood sexual abuse is associated with increased odds of using physical violence against people (91), a higher prevalence of any mental distress (90), but may be a risk factor for ‘sexual risk behaviours’ in persons with SMI (92, 93). Similarly, physical abuse in adulthood was a risk factor for psychiatric comorbidity, while sexual abuse in adulthood had a two-fold increased risk of having psychiatric comorbidity; findings are in agreement with previous studies which established that people with mental disabilities are subjected to abuse and neglect that exacerbates their illness (94-96). Results from this study indicate that mood stabilizers were associated with psychiatric comorbidity since they are commonly used to treat people with bipolar disorder, sometimes schizoaffective disorder and borderline personality disorder; in some cases, mood stabilizers are used to supplement other medications, such as antidepressants used to treat depression. However, a previous study suggests that psychotic features and other psychiatric disorders are associated with poor treatment response to mood stabilizers in some psychiatric patients (97). A previous study established that mood stabilizers, are associated with an increased risk for several physical diseases, including obesity, dyslipidemia, diabetes mellitus, thyroid disorders, hyponatremia; cardiovascular, respiratory tract, gastrointestinal, haematological, musculoskeletal and renal diseases, as well as movement and seizure disorders (98).

Among the psychiatric illness factors, having experienced a past psychotic episode was protective against HIV and the plausible explanation relates to the social stigma expressed towards the mentally sick people that could have limited their sexual involvement thus reduced risk for contracting HIV (84, 85).

Among the maladaptive behaviours, alcohol use had a two-fold increased risk of having syphilis; results from this study are in agreement with previous studies which suggest that there is a high correlation between alcohol use and sexually-transmitted diseases (STDs) because alcohol use strongly influences men and women to engage in risky activities while under the influence (99, 100). Alcohol subjects the user(s) into ‘a state of higher confidence and lower inhibition’, leading them to make riskier choices, like engaging in unprotected sexual activities (100).

Alcohol use was a risk factor for having hypertension; results from this study happen to be in agreement with findings from previous studies which established that alcohol use is widely recognized and a highly prevalent risk factor for hypertension which is supported by biochemical pathways (101, 102).

According to results from this study, mood stabilizers were associated with a two-fold increased risk of having obesity. Results from this study rhyme with findings established through review of studies which suggest that treatment with mood stabilisers is associated with overweight and obesity which results from excessive carbohydrate consumption, coupled with reduced physical exercises - common among patients with comorbidity (103). Alcohol drinking problems were associated with a ten-fold increased risk of obesity and the results are in agreement with a Ugandan study which suggests that frequent alcohol use (highly prevalent in Uganda) is a key risk factor for both hypertension and obesity (104). Similarly, use of marijuana was associated with eight times increased risk of having of obesity, contrary to findings from a study which suggests a lower prevalence of overweight and obesity among young adult cannabis users (105, 106); possible explanation could be that cannabis consumption reduces energy storage and increases metabolic rates, thus reversing the impact on body mass index of elevated dietary omega-6/omega-3 ratios (107). Use of khat was thirty-seven times associated with an increased risk of having obesity; evidence from recent review of the studies suggests that different khat extracts or cathinone produces changes in terms of weight, fat mass, appetite, lipid biochemistry and hormonal levels (108). The same review further suggests that the mechanism of these changes is the central action that produces changes in the physiology of dopamine and serotonin (108).

### Association between comorbid psychiatric and physical disorders and negative outcomes

In this study, lifetime suicide attempt was associated with a two-fold increased risk of having psychiatric comorbidity. Findings from this study were in agreement with a previous study which established that suicide risk is highly elevated among people with both physical and psychiatric illness (comorbidity) due to the double burden of both conditions (109). Similarly, risky sexual behaviour was associated with a two-fold increased risk of having psychiatric comorbidity. Findings from this study are in agreement with a previous study which suggests that there is a clear association between risky sexual behaviour and common psychiatric disorders; sexual risk taking behaviour is also increased with psychiatric comorbidity, possible due to the nature of the illness and loss of insight (110).

This study has many strong points such as use of an adequate sample size due to pooling of multiple longitudinal panels, ability to assess prevalence of physical and psychiatric comorbidity, clustering of physical and psychiatric disorders plus establishing factors associated with comorbid physical and psychiatric disorders. However, there are some limitations to consider when interpreting the findings. This was cross-sectional study therefore it was not possible to determine the causal pathway between physical and psychiatric comorbidity; thus a longitudinal study could be able to clearly establish such a causal pathway. Data on only a few physical morbidities were collected and considered in analysis; there is need to undertake a bigger study that focuses upon all physical and psychiatric comorbidities. Despite these limitations, to the best of our knowledge, this is the first study to establish the prevalence of physical and psychiatric comorbidity and associated risk factors among patients with severe mental illness attending Butabika National Psychiatric Referral Hospital and Masaka Regional Referral Hospital.

## Conclusion

Based upon the findings established by this study, prevalence of comorbidity and associated risk factors among patients with severe mental illness (SMI) in rural and urban Uganda has become increasingly imperative. This issue may be particularly important for psychiatric practice in Uganda, given that most people with SMI in Uganda experience higher levels of socio-economic deprivation which is strongly and consistent associated with comorbidity (111). Given the central role of mental illness within the comorbidity continuum, it is our debate that psychiatrists, physicians, Clinical Psychologists, psychiatric clinical officers (PCO’s), medical clinical officers (MCO’s), researchers and policy makers urgently need to discuss how best to develop and evaluate services that will improve physical, psychological and social outcomes for our patients. The emerging healthcare delivery models in Uganda should purpose to reduce fragmentation of care to help reduce the risks of preventable hospitalizations among mental healthcare beneficiaries with psychiatric comorbidity.

## Materials and methods

### Study design and site

A cross-sectional study was undertaken among 1,201 individuals with SMI attending care at the out-patient departments (OPDs) at Butabika hospitals (central) and Masaka hospital (southwestern) Uganda. To be eligible for the study, participants had to be adults over 18 years of age. Participants had to speak English or Luganda (the local language spoken in the study areas). Exclusion criteria were concurrent enrollment in another study), need of immediate medical attention, and unable to understand the study’s assessment instruments.

### Measures

The assessment battery comprised of a structured and standardised, locally translated psychosocial instruments (112-115). SMIs were established using the MINI International Neuropsychiatric Interview version 7.2. The tools were administered by trained psychiatric nurse / psychiatric clinical officer research assistants who assessed among others the psychiatric diagnosis. The variables reported in this paper include: (i) socio-demographic factors (study site, gender, age category, religion, socio-economic status, and marital status), (ii) psychosocial factors (social support, mental health stigma, childhood physical abuse, childhood sexual abuse, physical abuse in adulthood and sexual abuse in adulthood), (iii) psychiatric illness factors (family history of psychiatric illness, past depressive episode, past manic episode, past psychotic episode, lifetime suicide attempt), (iv) Psychotropic drugs (Antiparkinsonian medication, mood stabilizers, 1st generation neuroleptics, 2nd generation neuroleptics, Tri-cyclic anti-depressants, Selective Serotonin Reuptake Inhibitors) and (v) Maladaptive behaviour (alcohol use, use of tobacco, alcohol drinking problem, use of marijuana, use of khat).

### Statistical Analysis

Analyses were done using Stata release Stata 15 (StataCorp, TX, USA)

Factors associated with each morbidity were found by fitting separate logistic regression models. A binary variable “psychiatric comorbidity” was generated having two or more of current/past depression or current/past mania or current/past psychosis

Firstly, descriptive statistics were described for socio-demographic characteristics and psychosocial factors. All models included study site (as a design variable), sex and age as a priori confounders. Potential additional socio-demographic characteristics included social economic status (8 household item scale), marital status, employment status, religion and level of education.

The prevalence and patterns of psychiatric illness factors, maladaptive behavior characteristics and morbidities were investigated at 95% CI. Psychiatric and physical disorders were clustered to show different patterns of comorbidities.

Secondly, factors associated with having comorbid physical and psychiatric disorders among patients with severe mental illness were assessed by fitting multiple logistic regression models, adjusting for study site, age of the respondent and sex.

The impact of the morbidities with behavioral outcomes was investigated by fitting logistic regression models or ordinal logistic regression models. Logistic regression models were fitted for risky sexual behavior, symptom count on psychiatric outcomes and clinical outcomes (adherence to psychiatric medications and oral ART).

## Supporting information

Publications

Cover letter

## Data Availability

Data can be available from MRC repository upon request

## Declarations section

### Ethics approval and consent to participate

The study obtained ethical approvals from the Uganda Virus Research Institute’s Research and Ethics Committee (GC/127/19/10/612) and the Uganda National Council of Science and Technology (HS 2337). Participants were given information about the study by trained study psychiatric nurses and informed consent and assent sought before enrolment into the study. Participants found to have a SMI were provided healthcare and supported at the out-patient departments (OPDs) of their respective hospitals.

## Consent to publish

Not applicable.

## Availability of data and materials

All data and materials in this manuscript, additional files and figures attached are freely available with no restrictions.

## Authors’ contributions

RSM, WS, GZR, CB, KDG, AK, VP, MN and EK have made substantial contributions to conception, design, acquisition of data, drafting the manuscript, revising it critically and gave the final approval of this version to be published. WS did the analysis and interpretation of data.

Each author participated sufficiently in this work and takes public responsibility for appropriate portions of the content.

## Acknowledgments

The authors wish to thank the managers of the two study sites (Butabika National Psychiatric Referral Hospital and Masaka Regional Referral Hospital) for permitting the study to be conducted at their out-patient departments. The authors extend appreciation to the Medical Research Council, Uganda (MRC, Uganda) for funding and facilitating the study. Special gratitude is extended to the staff working at the two out-patient departments where the study was conducted. Appreciation is extended to the diligent work of research assistants. Gratitude is extended to the participants for their time and trust.

## Author’s information

^1^Mental Health Project, MRC/UVRI and LSHTM Uganda Research Unit, P. O. Box, 49, Entebbe, Uganda

^2^Butabika National Psychiatric Hospital, Kampala, Uganda

^8^Department of Mental health, School of Health Sciences, Soroti University, P. O. Box; 211, Soroti, Uganda

Tel: +256 772 592504, +256 446556 & +256 702 592504

E mail: &

---

**Figure 1:**
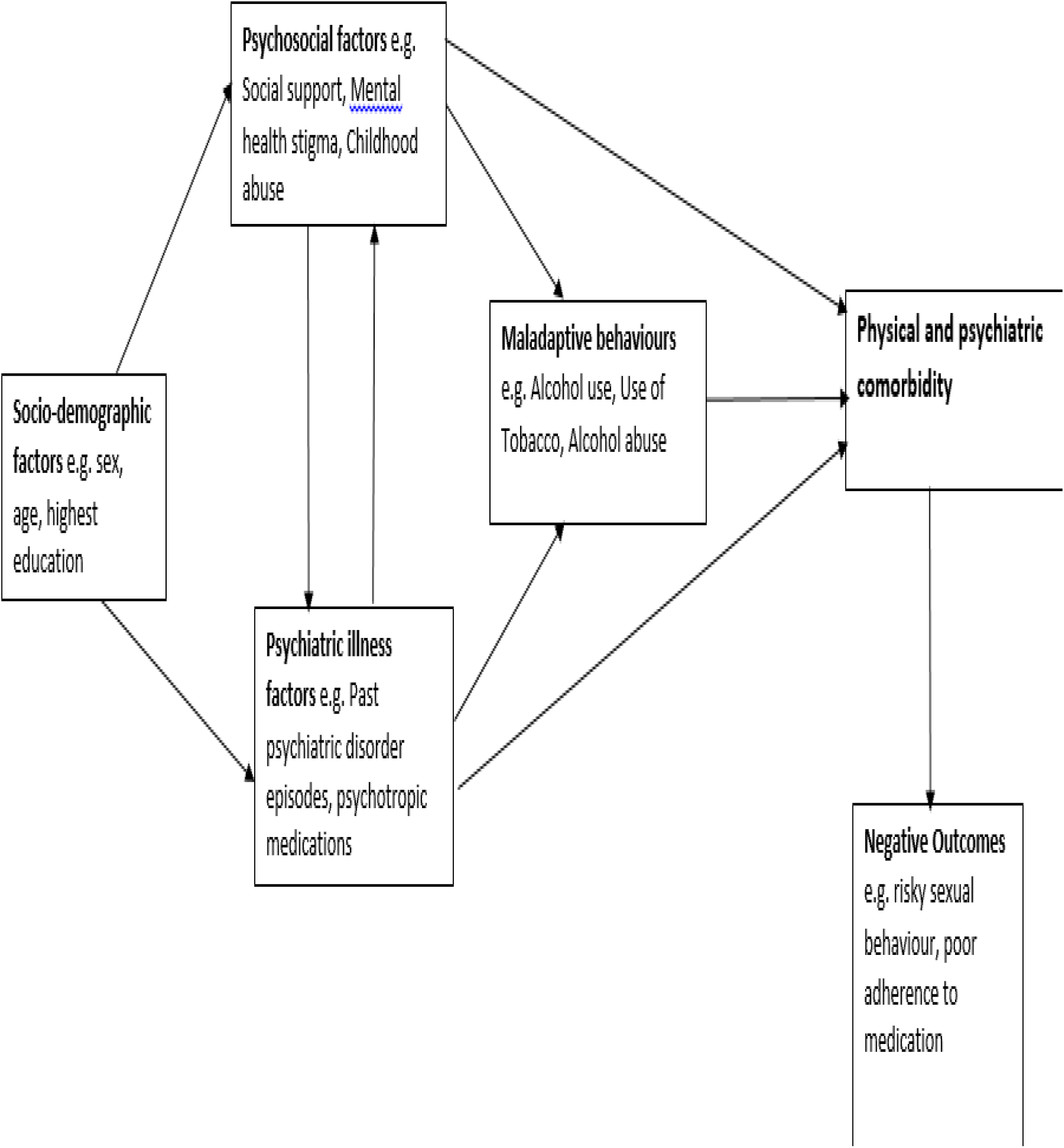
Conceptual framework on comorbidity in severe mental illness.

## Additional material

**Additional file 1:** Table 1: Socio demographic and psychosocial characteristics

**Additional file 2:** Table 3: Association between socio-demographic factors and comorbid psychiatric and physical disorders

**Additional file 3:** Table 4: Psychosocial and psychiatric factors associated with comorbid psychiatric and physical disorders

**Additional file 4:** Table 5: Association between comorbid psychiatric and physical disorders and negative outcomes

