## Supplemental Table 1 for "Physical and psychiatric comorbidity among patients with severe mental illness as seen in Uganda"

**Table 1: Socio demographic and psychosocial characteristics**

| Factor | **Frequency(n=1201)** | **(%)** |
| --- | --- | --- |
| **Site** | | |
| Butabika(urban) Masaka(rural) | 701  500 | 58.4%  41.6% |
| **Gender**  Male  Female | 547  654 | 45.5%  54.5% |
| **Age**  Mean(SD)  **Age**  <25  25 – 34  35 – 49  >=50 | 37.6(11.7)  138  384  475  200 | 11.5%  32.1%  39.7%  16.7% |
| **Socio-economic status(grouped)**  0 – 2  3 – 4  5 – 6  7 - 8 | 189  409  486  117 | 15.7%  34.1%  40.5%  9.7% |
| **Marital status**  Currently married  Widowed  Separated/divorced  Single | 384  57  295  464 | 32.0%  4.7%  24.6%  38.7% |
| **Employment status**  Farmer/fisherman  Professional  Informal employment  Unemployed | 304  139  206  549 | 25.4%  11.6%  17.2%  45.8% |
| **Religion**  Christian  Moslem  Others | 977  212  10 | 81.3%  17.6%  0.83% |
| **Education level**  No formal education  Primary  Secondary  Tertiary | 37  476  460 | 3.1%  39.6%  38.3%  22718.9% |
| **Psychosocial factors** |  |  |
| Childhood physical abuse | 395 | 32.9 % |
| Childhood sexual abuse | 296 | 24.7 % |
| Physical abuse in adulthood | 409 | 34.1% |
| Sexual abuse in adulthood | 263 | 21.9% |
| Family history of psychiatric illness | 731 | 61.0% |
