## Supplemental table 3 for "Physical and psychiatric comorbidity among patients with severe mental illness as seen in Uganda"

**Table 3: Association between socio-demographic factors and comorbid psychiatric and physical disorders**

| Factor | PSYCHIATRIC COMORBIDITY* | HIV* | SYPHILIS* | HYPERTENSION* | OBESITY* |
| --- | --- | --- | --- | --- | --- |
| Site  Butabika  Masaka | 1  0.77(0.60 ; 0.98)  **P = 0.034** | 1  1.11(0.71 ; 1.74)  P=0.639 | 1  0.74 (0.42 – 1.32)  P=0.312 | 1  0.64(0.49 ; 0.84)  **P=0.001** | 1  0.43(0.29 ; 0.62)  **P<0.001** |
| Sex  Male  Female | 1  1.07(0.84 ; 1.36)  P=0.570 | 1  2.62(1.57 ; 4.36)  **P<0.001** | 1  1.18 (1.02 ; 1.06)  P=0.551 | 1  0.92(0.71 ; 1.20)  P=0.547 | 1  4.66(3.05 ; 7.12)  **P<0.001** |
| Age category  18 – 24  25 – 34  35 – 49  >=50 | 1  1.10(0.73 ; 1.67)  0.94(0.63 ; 1.41)  0.61(0.39 ; 0.95)  **P=0.008** | 1  1.52(0.56 ; 4.15)  2.44(0.94 ; 6.36)  2.37(0.84 ; 6.66)  P=0.131 | 1  1.68(0.47 ; 5.96)  1.63(0.47 ; 5.71)  4.67(1.35 ; 16.20)  **P=0.003** | 1  1.30(0.76 ; 2.21)  2.49(1.50 ; 4.15)  3.79(1.50 ; 4.15)  **P<0.001** | 1  3.26(1.25 ; 8.50)  4.42(1.73 ; 11.24)  4.39(1.63 ; 11.74)  **P=0.011** |
| Religion  Christian  Moslem | 1  1.28(0.93 ; 1.77)  P=0.130 | 1  0.31(0.13 – 0.72)  **P=0.007** | 1  1.06(0.50 ; 2.23)  P=0.877 | 1  0.88(0.61 ; 1.25)  P=0.472 | 1  1.06(0.68 ; 1.67)  P=0.786 |
| SES index  Per unit increase | 0.97(0.90 ; 1.03)  P=0.327 | 0.87(0.77 ; 0.98)  **P=0.025** | 0.96(0.82 ; 1.13)  P=0.640 | 1.04(0.97 ; 1.12)  P=0.274 | 1.16(1.05 ; 1.28)  P**=0.003** |
| Marital status  Married  Widowed  Separated  Single | 1  0.71(0.39 ; 1.28)  1.08(0.78 ; 1.49)  0.77(0.57 ; 1.05)  P=0.136 | 1  2.95(1.26 ; 6.88)  1.30(0.73 ; 2.29)  0.72(0.38 ; 1.34)  **P=0.027** | 1  3.44(1.33 ; 8.94)  1.53(0.75 ; 3.17)  0.81(0.36 ; 1.85)  **P=0.044** | 1  1.58(0.87 ; 2.90)  1.03(0.73 ; 1.46)  0.98(0.69 ; 1.38)  P=0.503 | 1  1.17(0.56 ; 2.41)  0.80(0.52 ; 1.25)  0.67(0.43 ; 1.05)  P=0.286 |

**Note:** *Adjusted for study site, sex and age
