## Supplemental table 4 for "Physical and psychiatric comorbidity among patients with severe mental illness as seen in Uganda"

**Table 4: Psychosocial and psychiatric factors associated with comorbid psychiatric and physical disorders**

| Factor | PSYCHIATRIC COMORBIDITY* | | HIV* | SYPHILIS* | HYPERTENSION* | OBESITY* |
| --- | --- | --- | --- | --- | --- | --- |
| *Psychosocial factors* | | | | | | |
| Social support scale  Per unit increase | 0.98(0.96 ; 0.99)  **P=0.005** | 1.01(0.98 ; 1.03)  P=0.693 | | 1.01(0.97 ; 1.05)  P=0.563 | 0.99(0.98 ; 1.01)  P=0.446 | 0.99(0.97 ;1.01)  P=0.327 |
| Mental health stigma  Per unit increase | 1.04(1.03 ; 1.06)  **P<0.001** | 1.01(0.98 ; 1.04)  P=0.666 | | 0.99(0.96 ; 1.03)  P=0.656 | 0.99(0.98 ; 1.01)  P=0.753 | 0.99(0.97 ; 1.01)  P=0.280 |
| Childhood physical abuse  Per unit increase | 1.07(1.03 ; 1.10)  **P<0.001** | 0.99(0.93 ; 1.06)  P=0.856 | | 1.00(0.93 ; 1.08)  P=0.974 | 0.99(0.96 ; 1.03)  P=0.883 | 0.97(0.93 ; 1.02)  P=0.286 |
| Childhood sexual abuse  Per unit increase | 1.06(1.03 ; 1.10)  **P=0.001** | 1.03(0.98 ; 1.08)  P=0.227 | | 1.03(0.96 ; 1.09)  P=0.430 | 1.01(0.98 ; 1.05)  P=0.457 | 0.99(0.96 ; 1.04)  P=0.916 |
| Physical abuse in adulthood  Yes | 1.69(1.30 ; 2.20)  **P<0.001** | 1.36(0.86 ; 2.16)  P=0.186 | | 1.13(0.63 ; 2.02)  P=0.689 | 1.20(0.91 ; 1.57)  P=0.192 | 1.03(0.72 ; 1.48)  P=0.871 |
| Sexual abuse in adulthood  Yes | 2.22(1.60 ; 3.08)  **P<0.001** | 1.57(0.95 ; 2.58)  P=0.077 | | 1.00(0.50 ; 1.99)  P=0.991 | 1.22(0.89 ; 1.69)  P=0.216 | 0.97(0.65 ; 1.43)  P=0.872 |
| *Psychiatric illness factors* | | | | | | |
| Family history of psychiatric illness – yes | 1.24(0.97 ; 1.58)  P=0.088 | 0.80(0.51 ; 1.25)  P=0.325 | | 0.86(0.49 ; 1.52)  P=0.610 | 1.11(0.84 ; 1.45)  P=0.458 | 1.07(0.75 ; 1.53)  P=0.692 |
| Past depressive episode  Yes | - | 1.02(0.62 ; 1.68)  P=0.940 | | 0.98(0.54 ; 1.79)  P=0.959 | 0.94(0.70 ; 1.26)  P=0.695 | 1.37(0.93 ; 2.00)  P=0.110 |
| Past manic episode  Yes | - | 1.20(0.73 ; 1.97)  P=0.474 | | 1.33(0.70 ; 2.52)  P=0.377 | 1.07(0.80 ; 1.43)  P=0.635 | 1.40(0.94 ; 2.08)  P=0.096 |
| Past psychotic episode  Yes | - | 0.59(0.37 ; 0.94)  **P=0.027** | | 0.97(0.51 ; 1.85)  P=0.932 | 1.09(0.81 ; 1.50)  P=0.554 | 0.80(0.54 ; 1.18)  P=0.263 |
| *Psychotropic drugs* | | | | | | |
| Antiparkinsonian medication  Yes | 1.36(0.87,2.13)  P=0.176 | 0.66(0.31,1.38)  P=0.268 | | 2.07(0.49,8.80)  P=0.323 | 0.74(0.46,1.21)  P=0.234 | 1.96(0.87,4.41)  P=0.106 |
| Mood stabilizers  Yes | 1.43(1.12,1.82)  **P=0.004** | 1.07(0.68,1.69)  P=0.763 | | 0.71(0.40,1.25)  P=0.234 | 1.17(0.90,1.53)  P=0.238 | 1.62(1.14,2.31)  **P=0.008** |
| 1^st^ generation neuroleptics  Yes | 1.39(0.94,2.06)  P=0.097 | 1.00(0.48,2.08)  P=0.993 | | 0.83(0.34,2.02)  P=0.685 | 0.88(0.57,1.36)  P=0.559 | 1.39(0.76,2.55)  P=0.281 |
| 2^nd^ generation neuroleptic  Yes | 0.78(0.49,1.25)  P=0.300 | 1.19(0.51,2.75)  P=0.693 | | 0.52(0.12,2.23)  P=0.380 | 1.18(0.71,1.94)  P=0.522 | 1.10(0.60,2.01)  P=0.767 |
| Tri-cyclic anti-depressants  Yes | 0.84(0.57,1.24)  P=0.377 | 0.81(0.39,1.69)  P=0.573 | | 0.74(0.28,1.94)  P=0.539 | 1.11(0.72,1.69)  P=0.641 | 1.08(0.64,1.81)  P=0.769 |
| SSRIs  Yes | 1.21(0.76,1.91)  P=0.422 | 0.82(0.34,1.95)  P=0.653 | | 1.98(0.85,4.59)  P=0.111 | 1.09(0.67,1.78)  P=0.715 | 0.83(0.43,1.59)  P=0.566 |
| *Maladaptive behaviour* | | | | | | |
| Alcohol use  Yes | 1.36(0.92 ; 2.10)  P=0.121 | 1.04(0.52 ; 2.10)  P=0.893 | | 2.37(1.19 ; 4.70)  **P=0.013** | 1.52(1.03 ; 2.26)  **P=0.037** | 0.96(0.53; 1.71)  P=0.893 |
| Use of Tobacco  Yes | 0.99(0.59 ; 1.66)  P=0.986 | 0.25(0.03 ; 1.86)  P=0.177 | | 0.96(0.28 ; 3.25)  P=0.950 | 1.12(0.64 ; 1.95)  P=0.673 | 0.89(0.36; 2.18)  P=0.797 |
| Alcohol drinking problem (CAGE positive) – yes | 0.69(0.14 ; 3.52)  P=0.664 | 0.67(0.06 ; 7.08)  P=0.740 | | 4.69(0.54 ; 40.64)  P=0.161 | 1.76(0.46 ; 6.73)  P=0.406 | 9.90(1.20; 81.5)  **P=0.033** |
| Use of marijuana  Yes | 0.80(0.17 ; 3.63)  P=0.769 | - | | - | 3.45(0.72 ; 16.39)  P=0.119 | 7.62(1.31; 44.25)  **P=0.024** |
| Use of Khat  Yes | 1.09(0.20 ; 6.03)  P=0.922 | - | | - | 0.74(0.08 ; 6.60)  P=0.788 | 37.05(6.09; 225.6)  **P<0.001** |

**Note:** *Adjusted for study site, sex and age; SSRIs- Selective Serotonin Reuptake Inhibitors
