## Supplemental table 5 for "Physical and psychiatric comorbidity among patients with severe mental illness as seen in Uganda"

**Table 5 Association between comorbid psychiatric and physical disorders and negative outcomes**

| Factor | PSYCHIATRIC COMORBIDITY* | HIV* | SYPHILIS* | HYPERTENSION* | OBESITY* |
| --- | --- | --- | --- | --- | --- |
| Behavioural outcomes | | | | | |
| Lifetime suicide attempt – yes | 1.99(1.44 ; 2.74)  **P<0.001** | 1.05(0.61 ; 1.82)  P=0.857 | 0.94(0.46 ; 1.92)  P=0.878 | 0.76(0.55 ; 1.07)  P=0.121 | 1.26(0.83 ; 1.91)  P=0.271 |
| Risky sexual behaviour – yes | 1.77(1.28 ; 2.30)  **P<0.001** | 1.46(0.88; 2.40)  P=0.140 | 1.44(0.77 ; 2.70)  P=0.250 | 0.89(0.65; 1.22)  P=0.469 | 0.75(0.49; 1.15)  P=0.186 |
| Clinical outcomes |  |  |  |  |  |
| Missed taking psychiatric medications - yes | 1.01(0.75; 1.36)  P=0.945 | 0.81(0.46;1.43)  P=0.460 | 0.85(0.41 ; 1.73)  P=0.649 | 0.79(0.57; 1.09)  P=0.156 | 0.78(0.51; 1.21)  P=0.271 |
| Missed taking oral ART – yes | 1.61(0.28; 9.13)  P=0.589 | 0.13(0.01; 2.35)  P=0.169 | 3.86(0.49 ; 30.3)  P=0.199 | 1.49(0.22; 10.02)  P=0.684 | - |
