## Supplementary material for "Physical and psychiatric comorbidity among patients with severe mental illness as seen in Uganda": Publications

**Publications** ;Grouped by year of publications (starting from the most recent)

**2020**

1. Prevalence and risk factors for youth suicidality among perinatally infected youths living with HIV/AIDS in Uganda: the CHAKA study

Godfrey Zari Rukundo, **Richard Stephen Mpango**, Wilber Ssembajjwe, Kenneth D. Gadow,

, Vikram Patel and Eugene Kinyanda

Published by Child and Adolescent Psychiatry and Mental Health, 24th.October 2020

<https://doi.org/10.1186/s13034-020-00348-0>

<https://rdcu.be/b87jA>

2. Anxiety disorders and asthma among adolescents in urban Uganda: the role of early life exposures

Harriet Mpairwe, **Richard Stephen Mpango**, Wilber Sembajjwe, Emily L Webb, Alison M Elliott, Neil Pearce, Eugene Kinyanda

Preprint for European Respiratory journal, 14 October 2020

<https://doi.org/10.1101/2020.10.08.20209478>

3. Challenges to peer support in low- and middle-income countries during COVID-19

**Richard Mpango**, Jasmine Kalha, Donat Shamba, Mary Ramesh, Fileuka Ngakongwa, Arti Kulkarni, Palak Korde, Juliet Nakku and Grace K. Ryan

Published by Globalization and Health, 25 September 2020

<https://doi.org/10.1186/s12992-020-00622-y>

<https://rdcu.be/b7JvQ>

4. Peer support for people with severe mental illness versus usual care in high-, middle- and low-income countries: study protocol for a pragmatic, multicentre, randomised controlled trial (UPSIDES-RCT)

Galia S. Moran, Jasmine Kalha, Annabel S. Mueller-Stierlin, Reinhold Kilian, Silvia Krumm, Mike Slade, Ashleigh Charles, Candelaria Mahlke, Rebecca Nixdorf, David Basangwa, Juliet Nakku,

**Richard Mpango**, Grace Ryan, Donat Shamba, Mary Ramesh, Fileuka Ngakongwa, Alina Grayzman,

Soumitra Pathare, Benjamin Mayer & Bernd Puschner

Published by Trials BMC, volume 21, Article number: 371 (2020),May 2020

https://doi.org/10.1186/s13063-020-4177-7

5. Clinical correlates and adverse outcomes of ADHD, disruptive behavior disorder and their co-occurrence among children and adolescents with HIV in Uganda

Tatiana Taylor Salisbury, Eugene Kinyanda, Jonathan Levin, Alexander Foster, **Richard Mpango**, Vikram Patel & Kenneth D. Gadow

Accepted for publication by Journal of AIDS Care, March 2020

6. Major depressive disorder among HIV infected youth in Uganda: Incidence, persistence and their predictors

Eugene Kinyanda, Tatiana T. Salisbury, Sylvia Kiwuwa Muyingo, Wilber Ssembajjwe, Jonathan Levin, Noeline Nakasujja, **Richard S. Mpango**, Catherine Abbo, Soraya Seedat, Ricardo Araya, Seggane Musisi, Kenneth D. Gadow, Vikram Patel

Published by the journal AIDS and Behaviour, February 2020

7. Peer support for people with severe mental illness versus usual care: study protocol for a pragmatic multicentre randomised controlled trial (UPSIDES-RCT)

Galia Sharon Moran, Jasmine Kalha, Annabel Mueller-Stierlin, Reinhold Kilian, Silvia Krumm, Mike Slade, Ashleigh Charles, Candelaria Mahlke, Rebecca Rebecca Nixdorf, David Basangwa, Juliet Nakku, **Richard Mpango**, Grace Ryan, Donat Shamba, Mary Ramesh, Fileuka Ngakongwa, Alina Grayzman, Soumitra Pathare, Benjamin Mayer, Bernd Puschner

Published by Integrative & Complementary Medicine - Internal Medicine, Trials BMC, 13^th^.February 2020

10.21203/rs.2.20188/v1

8. Substance use among HIV-infected adolescents in Uganda: rates and association with potential risks and outcome factors

C. Birungi, W. Ssembajjwe, T. T. Salisbury, J. Levin, N. Nakasujja, R. S. **Mpango**, C. Abbo, S. Seedat, R. Araya, S. Musisi, K. D. Gadow, V. Patel & E. Kinyanda

Published by the journal of AIDS Care , January 2020

10.1080/09540121.2020.1717419

9. Typology of modifications to peer support work for adults with mental health problems: systematic review

Ashleigh Charles, Dean Thompson, Rebecca Nixdorf, Grace Ryan, Donat Shamba, Jasmine Kalha, Galia Moran, Ramona Hiltensperger, Candelaria Mahlke, Bernd Puschner, Julie Repper, Mike Slade, **Richard Mpango**

Published by the British Journal of Psychiatry, January 2020

<https://www.cambridge.org/core/journals/the-british-journal-of-psychiatry/article/typology-of-modifications-to-peer-support-work-for-adults-with-mental-health-problems-systematic-review/E3410880C3F33A8478E570AAD0A0EE15>

10.1192/bjp.2019.264

**2019**

1. Adaptation and validation of a brief DSM-5 based psychiatric rating scale for childhood and adolescent mental health in Uganda: The Child and Adolescent Symptom Inventory-Progress Monitor (CASI-PM)

**Richard Stephen Mpango**, Wilber Ssembajjwe, Sylvia Kiwuwa Muyingo, Kenneth D. Gadow, Vikram Patel, Eugene Kinyanda

Published by the journal of Vulnerable Children and Youth Studies; Journal Subtitle- An International Interdisciplinary Journal for Research, Policy and Care, October 2019

10.1080/17450128.2019.1686672

2. Peer Support for Frequent Users of Inpatient Mental Health Care in Uganda: Protocol of a Quasi-Experimental Study

Grace K. Ryan, Mauricia Kamuhiirwa, James Mugisha, Dave Baillie, Cerdic Hall, Carter Newman, Eddie Nkurunungi, Sujit D. Rathod, Karen M. Devries, Mary J. De Silva, **Richard Mpango**

Published by the BMC psychiatry, November 2019

3. Agreement and Discrepancy on Emotional and Behavioral Problems between Caregivers and HIV Infected Children and Adolescents from Uganda

Leigh Luella Van Den Heuvel, Jonathan Levin, **Richard Stephen Mpango**, Kenneth D Gadow, Vikram Patel, Jean B Nachega, Soraya Seedat, Eugene Kinyanda

Published by the journal of Frontiers in Psychiatry, section Child and Adolescent Psychiatry, June 2019

**https://www.frontiersin.org**

4. A systematic review of influences on implementation of peer support work for adults with mental health problems

Nashwa Ibrahim, Dean Thompson, Rebecca Nixdorf, Jasmine Kalha, **Richard Mpango**, Galia Moran, Annabel Mueller‑Stierlin, Grace Ryan, Candelaria Mahlke, Donat Shamba, Bernd Puschner, Julie Repper, Mike Slade

Published by the journal of Social Psychiatry and Psychiatric Epidemiology, June 2019

<https://doi.org/10.1007/s00127-019-01739-1>

5. Service user involvement in global mental health: what have we learned from recent research in low and middle-income countries?

Grace K. Ryan, Maya Semrau, Eddie Nkurunungi, and **Richard S. Mpango**

Published by Current Opinion in Psychiatry, Lancet, April 2019

<http://dx.doi.org/10.1097/YCO.0000000000000506>

6. Rates, types and co-occurrence of emotional and behavioural disorders among perinatally HIV infected youth in Uganda: the CHAKA Study.

Eugene Kinyanda, Tatiana Taylor Salisbury, Jonathan Levin, Noeline Nakasujja, Richard Mpango, Catherine Abbo, Soraya Seedat, Ricardo Araya, Seggane Musisi, Kenneth D. Gadow, Vikram Patel

Published by the journal of Social psychiatry and psychiatric epidemiology , April 2019

**10.1007/s00127-019-01675-0**

7. The Burden and Risk Factors for Postnatal Depression and Depressive Symptomatology among Women in Kampala

Margaret Nampijja, Barnabas Natamba, **Richard Mpango** & Eugene Kinyanda

Published by the Tropical Doctor, February 2019.

[10.1177/0049475519837107](https://doi.org/10.1177/0049475519837107)

8. Prevalence, correlates for early neurological disorders and association with functioning among children and adolescents with HIV/AIDS in Uganda.

**Richard Stephen Mpango**, Godfrey Zari Rukundo, Sylvia Kiwuwa Muyingo^,^ Kenneth D. Gadow, Vikram Patel, Eugene Kinyanda

Published by BMC Psychiatry, 23rd.January 2019

<https://doi.org/10.1186/s12888-019-2023-9>

**2018**

1. Major Depressive Disorder: Longitudinal Analysis of Impact on Clinical and Behavioral Outcomes in Uganda

Eugene Kinyanda, Jonathan Levin, Noeline Nakasujja, Harriet Birabwa, Juliet Nakku, **Richard Mpango**, Heiner Grosskurth, Soraya Seedat, Ricardo Araya, Maryam Shahmanesh, and Vikram Patel

Published by the Journal of Acquired Immune Deficiency Syndromes (JAIDS) 78(2):1, February 2018

https://doi.org/10.1097/QAI.0000000000001647

2.Influences on implementation of peer support work for adults with mental health problems: A rapid review

Jasmine Kalha, **Richard Mpango**, Galia Moran, Ben Gurion,, Annabel Mueller-Stierlin, Grace Ryan, Candelaria Mahlke, Rebecca Nixdorf, Bernd Puschner, Julie Repper, Mike Slade

Published by PROSPERO; International prospective register of systematic reviews (NHS**),** July 2018

**https://doi.org/10.1007/s00127-019-01739-1**

3.A systematic review of Peer Support Worker training for adults with mental health problems

Candelaria Mahlke, Rebecca Nixdorf, Jasmine Kalha, **Richard Mpango**, Galia Moran, Annabel Mueller-Stierlin, Grace Ryan, Thomas Becker, Julie Repper, Dean Thompson, Mike Slade

Published by the PROSPERO; International prospective register of systematic reviews (NHS), August 2018

[**https://www.crd.york.ac.uk/prospero/display_record.php?RecordID=107772**](https://www.crd.york.ac.uk/prospero/display_record.php?RecordID=107772)

4.Exploration of the Understanding and etiology of ADHD in HIV/AIDS as observed by Adolescents with HIV/AIDS, Caregivers and Health Workers - using case vignettes

**Mpango, S. R**, Kinyanda,E., Rukundo,G.Z., Levin,J., Osafo,J., Gadow,D.K.

Published by the journal of African Health Sciences, August 2018 - Vol 18, No 3 (2018)

ScholarOne, 375 Greenbrier Drive, Charlottesville, VA, 22901

[**https://www.ajol.info/index.php/ahs/issue/view/17224**](https://www.ajol.info/index.php/ahs/issue/view/17224)

5.Caregiver and youth self-reported emotional and behavioural problems in Ugandan HIV-infected children and adolescents

Leigh L. van den Heuvel, Jonathan Levin, **Richard S. Mpango**, Kenneth D. Gadow, Vikram Patel, Jean B. Nachega, Soraya Seedat, Eugene Kinyanda

Published by the South African Journal of Psychiatry , 20 September 2018 | Vol 24 | a1265 |

[**https://sajp.org.za/index.php/sajp/article/view/1265**](https://sajp.org.za/index.php/sajp/article/view/1265)

https://doi.org/10.4102/sajpsychiatry.v24i0.1265

**2017**

1. Major depressive disorder and suicidality in early HIV infection and its association with risk factors and negative outcomes as seen in semi-urban and rural Uganda

Eugene Kinyanda, Noeline Nakasujja, Jonathan Levin, Harriet Birabwa, **Richard Mpango**, Heiner Grosskurth, Soraya Seedat, Vikram Patel

Published by the Journal of Affective Disorders , 23 January 2017

**https://dx.doi.org/10.1016/j.jad.2017.01.033**

2. Incidence and persistence of major depressive disorder among people living with HIV in Uganda

Eugene Kinyanda, Helen A. Weiss, Jonathan Levin, Noeline Nakasujja, Harriet Birabwa, Juliet Nakku, **Richard Mpango**, Heiner Grosskurth, Soraya Seedat, Ricardo Araya, Vikram Patel

Published by the Journal of AIDS Behaviour, June 2017 ; 21(6):1641-1654.

**https://doi.org/10.1007/s10461-016-1575-7.**

PMID: 27722834

3. Cross-cultural adaptation of the Child and Adolescent Symptom Inventory-5 (CASI-5) for use in central and south-western Uganda: the CHAKA project.

**Mpango, S. R.**, Kinyanda,E., Rukundo,G.Z., Levin,J., Gadow,D.K.

First Published by the Tropical Doctor, August 2, 2017 Research Article [**http://journals.sagepub.com/eprint/afJwfVX4jVqeQdS4xT5V/full**](http://journals.sagepub.com/eprint/afJwfVX4jVqeQdS4xT5V/full)**.**

4. Prevalence and correlates for ADHD and relation with social and academic functioning among children and adolescents with HIV/AIDS in Uganda

**Mpango, S**. R, Kinyanda,E., Rukundo,G.Z., Levin,J., Gadow,D.K.

Published by BMC Psychiatry, 26^th^.September 2017

Manuscript Number: BPSY-D-17-00418R1

[**https://bmcpsychiatry.biomedcentral.com/articles/10.1186/s12888-017-1488-7**](https://bmcpsychiatry.biomedcentral.com/articles/10.1186/s12888-017-1488-7)

**2016**

**2015**

Diaspora and peer support working: benefits of and challenges for the Butabika– East London Link

Dave Baillie, Mariam Aligawesa, Harriet Birabwa-Oketcho, Cerdic Hall, David Kyaligonza, **Richard Mpango**, Moses Mulimira and Jed Boardman

Published by BJPsych International, 2015

**2014**

1. Mental Health Practitioners' Reflections on Psychological Work in Uganda; Contrasting Perspectives from Two Professions

Jennifer Hall, Patricia d’Ardenne, James Nesereko, Roscoe Kasujja, David Baillie, **Richard Mpango** & Harriet Birabwa

Published by British Journal of Guidance and Counselling, February 2014

2. Psychiatric morbidity in early HIV infection and its association with risk factors and negative outcomes as seen in urban and rural Uganda

Eugene Kinyanda, Jonathan Levin, Noeline Nakasujja, Harriet Birabwa, Juliet Nakku, **Richard Mpango**, Heiner Grosskurth, Soraya Seedat, Vikram Patel

Published in 2014

**2013**

Symposium; An African Approach for training, supervising and practising CBT Symposium -Convened by

Elaine Hunter, David Baillie, James Nsereko, Rosco Kasujja, **Richard Mpango**, Dorothy Kiiza and Harriet Birabwa

Published by EABABCP, 2013
