## Supplementary material for "Physical and psychiatric comorbidity among patients with severe mental illness as seen in Uganda": Cover letter

10^th^.February, 2020

**The Editor-in-Chief,**

**eLife**

Dear Sir/Madam

,

**Re:** Initial submission of manuscript on**: ‘Physical and psychiatric comorbidity among patients with severe mental illness as seen Uganda’**

Mental ill health is a major cause of morbidity and mortality globally, with low- and middle-income countries (LMIC) experiencing a rapidly increasing prevalence (World Health Organization, 2017). Patients with severe mental illness (SMI) such as schizophrenia, depression and bipolar affective disorder die younger compared to other people in the general population (Crump C, Winkleby MA, Sundquist K, Sundquist J., 2013; Henderson M, Hotopf M, Shah I, Hayes RD, Kuh D., 2011; Lawrence D, Hancock KJ, Kisely S., 2013). However, most of the increased mortality among people with SMI is attributed to comorbidity, particularly with other non-communicable diseases (NCDs) (Crump C, Winkleby MA, Sundquist K, Sundquist J., 2013; Henderson M, Hotopf M, Shah I, Hayes RD, Kuh D., 2011; Laursen TM, Munk-Olsen T, Vestergaard M., 2012). Although comorbidity is a worldwide concern, the majority of the evidence comes from high-income countries, with few studies from LMIC, particularly sub-Saharan Africa (World Health Organization, 2017; Gladstone M, Lancaster GA, Umar E, Nyirenda M, Kayira E, van den Broek NR, et al., 2010). The purpose of this current study is to establish the prevalence of comorbidity and associated risk factors among patients with severe mental illness in rural and urban Uganda.

This work will contribute to the literature and evidence of the increasing burden of ‘physical and psychiatric comorbidity among patients with severe mental illness in Africa’; in Africa, detection and management of mental and physical comorbidity poses a particular challenge for LMIC healthcare systems with separate services for mental and physical health (World Health Organisation, 2017; Ajmera M, Wilkins TL, Findley PA, Sambamoorthi U., 2012). This work will cause debate among practitioners, researchers and policy makers to discuss how best to develop and evaluate services that will improve physical, psychological and social outcomes for our patients and lay a platform to establish interventions for ‘physical and psychiatric comorbidity among patients with severe mental illness in Africa’.

Information in this paper will be useful to practitioners (psychiatrists, physicians, clinical psychologists, clinical officers, nurses, etc.), researchers and policy makers interested in the increasing burden of ‘physical and psychiatric comorbidity among patients with severe mental illnesses.

This study has many strong points such as use of an adequate sample size due to pooling of multiple longitudinal panels, ability to assess prevalence of psychiatric comorbidity, clustering of physical and psychiatric disorders plus establishing factors associated with comorbid physical and psychiatric disorders. However, there are some limitations to consider when interpreting the findings. This was cross-sectional study therefore it was not possible to determine the causal pathway between physical and psychiatric comorbidity; thus a longitudinal study could be able to clearly establish such a causal pathway. Data on only a few physical morbidities were collected and considered in analysis; there is need to undertake a bigger study that focuses upon all physical and psychiatric comorbidities. Despite these limitations, to the best of our knowledge, this is the first study to establish the prevalence of physical and psychiatric comorbidity and associated risk factors among patients with severe mental illness attending Butabika national referral mental hospital and Masaka regional referral hospital.

I will appreciate when this work is considered for publication by your journal

Yours Faithfully,

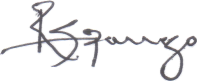

**Dr. Richard S. Mpango, PhD**

**Scientist / Project Leader, Mental Health Project,**

**MRC/UVRI and LSHTM Uganda Research Unit (MUL)**
